# Epidemiology and determinants of vitamin D deficiency in Eastern Nepal: a community-based, cross-sectional study

**DOI:** 10.1101/2022.05.10.22274787

**Authors:** O Sherchand, J K Baranwal, B Gelal

**Affiliations:** Department of Biochemistry, Nepal Medical College and Teaching hospital, Kathmandu, Nepal; Department of Biochemistry, B.P. Koirala Institute of Health Sciences, Dharan, Nepal

**Keywords:** serum 25(OH) vitamin D, risk factors, eastern Nepal

## Abstract

**Objective:** To estimate the prevalence of vitamin D deficiency in the Eastern the part of Nepal and identify the sociodemographic factors associated with it.

**Methods:** A descriptive cross-sectional study was carried out among 324 participants between the ages of 18 to 65 years from the Sunsari and Morang districts of Nepal. A semi-structured questionnaire helped obtain sociodemographic data followed by anthropometric measurements and blood sampling. 25(OH)D level was measured by Chemiluminescence immunoassay (CLIA) via a fully automated Maglumi 1000 analyzer (SNIBE Co, Ltd, China). Serum 25(OH)D was classified as deficient, insufficient, and sufficient (<20 ng/ml, 20-29 ng/ml, and 30–100 ng/ml respectively). The Chi-square test was used to analyze the sociodemographic variables followed by a post-hoc analysis. Significant variables were subject to multivariate logistic regression.

**Result:** 181(55.9%) of the study population had vitamin D deficiency. There was significant association between vitamin D status and time of maximum sun exposure (Chi-sq = 11.1, p=0.02), duration of sun exposure (Chi-sq = 15.1, p=0.004), type of meat intake (Fischer’s exact test is 16.4, p=0.01), frequency of fish intake (Fischer’s exact test is 19.3, p=0.001), frequency of dairy intake (Chi-sq=11.2, p=0.02). In multivariate regression, consumption of dairy products ≥3/week had lower OR (95% CI) [0.3(0.1-0.8) p:0.02] and weekly fish consumption had lower OR (95% CI) [0.06(0.008-0.6) p: 0.01] for vitamin D deficiency.

**Conclusion:** The prevalence of vitamin D deficiency was relatively high in eastern Nepal. This highlights the need to create public awareness regarding the importance of bare skin sun exposure and other sources of vitamin D as well as the need to implement food fortification policies by the government.

## 1. Background

In the recent years, hospitals across Nepal have witnessed an upsurge in vitamin D deficiency cases. Hospital-based studies conducted in Western and Southern Nepal depicted a high prevalence of vitamin D deficiency. [1, 2] However, data from the eastern parts is lacking which has warranted the need to determine the prevalence of vitamin D deficiency in this region.

Vitamin D deficiency is a pandemic health problem with multiple adverse health consequences. [3-5] Vitamin D is synthesized in the skin when exposed to ultraviolet (UV rays) from the sun. [5] Factors compromising the intensity of UV rays such as air pollution, latitude of residence, and skin pigmentation can impact vitamin D synthesis. [6-8] Another important source of vitamin D are dietary sources such as fish, cod liver oil, egg yolk, and mushroom; but the affordability of these items is beyond the reach of the majority of the households in Nepal. [9, 10]

Data from the neighbouring countries of Nepal; India and China both report a high prevalence of vitamin D deficiency. [11,12] However, there is a scarcity of data from Nepal as stipulated by a meta-analysis examining the prevalence of vitamin D deficiency in South Asians; which uncovered only two studies from Nepal. [13] Moreover, studies done in Nepal are hospital-based which can be subjected to clinical conditions that affect vitamin D metabolism, thus not representative of the community.

In this context, we conducted a community-based study to determine the prevalence of vitamin D deficiency and examined the different facets of vitamin D deficiency in terms of demographics, socioeconomic status, sun exposure factors, dietary and lifestyle pattern.

## 2. Methods

### 2.1. Study Design, Site and Participants

We conducted a community-based cross-sectional study in Sunsari and Morang districts (26°N) of eastern Nepal, between the months of April 2019 to March 2020. The sample size was calculated using the formula: n=z^2^ pq/d^2^

Where n=sample size, z= z score at 95% confidence interval (1.96)

P=estimated prevalence of vitamin d deficiency as 73.68% [2]

q=1-p, d=margin of error (0.05)

n = 296

We selected Belbari municipality, a major suburb of Morang district, and Dharan and Itahari, sub-metropolitan cities from Sunsari. With the aid of a community health worker, we recruited 337 participants; 225 from Morang and 112 from Sunsari. People were explained the purpose of our visit and those giving written informed consent were enrolled. We excluded pregnant women, lactating mothers and people taking vitamin D supplements, suffering from skin, liver and kidney diseases. Following the exclusion criteria; 10 participants from Sunsari and 3 from Morang were excluded. This study was initiated after receiving ethical approval from the Institutional Review Committee, B.P Koirala Institute of Health Sciences.

### 2.2. Demographic variables

We administered a semi-structured questionnaire to obtain demographic data [age: young age (18-44 years) and middle aged and elderly (45-65 years), gender (male and female), ethnicity (Brahmin and Chettri, Newar, Janajati, occupational caste), highest education level attained (upto primary school, intermediate, high school, above high school), occupation (professional and semi-professionals, skilled and semi-skilled work, arithmetic skill jobs, unskilled work, unemployed)]. An aggregate of educational status, occupation of the head of the household and monthly income of the household helped determine socioeconomic status. [14] Smoking status was classified as current smoker, former or never smoker. Alcohol intake was divided into drinks alcoholic beverage, does not drink. Questionnaire regarding indicators of sun exposure included; duration of sun exposure (<15 minutes, 15-30 minutes and >30 minutes), time of the day during maximum sun exposure (early morning 6-8:59 am, late morning 9-11:59 am and afternoon 12 noon to evening), skin colour (fair, light brown, dark brown), sunscreen (uses SPF 15 and above, does not use). Physical activity was classified as active if they exercised >30 minutes/day at least 5 days a week and moderate if they exercised but lesser than the first criteria and sedentary if they had no physical activity or irregular activity. Body mass index (BMI) was calculated as weight by height squared (kg/meter^2^) and classified as normal weight (18.5–22.9 kg/m^2^), overweight (≥23.0–24.99 kg/m^2^), and obese (≥25 kg/m^2^)] [15]

### 2.3. Dietary pattern

The participants were asked if they followed a vegetarian or non-vegetarian diet. If non-vegetarian; the type of meat consumed: a. only chicken, b. chicken and mutton, c. chicken and pork or d. none. Food frequency was enquired by asking “how often they consumed: meat: (a. ≥2/week, b. once a week, c. 2-3 times/month d. none), milk (a. ≥3/week, b. 1-2/week, c. less than once a week), dairy products as curd, cheese, butter, ghee, including egg: (a. ≥3/week, b. 1-2/week, c. less than once a week), fish: (a. monthly, b. weekly, c. rarely/never).

### 2.4. Serum 25(OH)D measurement

1,25(OH)D is the active form of vitamin D however, studies have identified serum 25(OH)D as the best marker of vitamin D status. [16] We used the same to find the vitamin D status. Venous blood samples were collected in a 3 ml plain vial and centrifuged to separate serum. Aliquots of serum samples were transported to the biochemistry laboratory of BPKIHS maintaining a cold chain and stored at -20° C for subsequent batch analysis of serum 25(OH)D. We measured serum 25(OH)D with Maglumi 1000 analyzer with Chemiluminescence Immunoassay (SNIBE Co, Ltd, China). Quality was assured using internal quality control provided by the manufacturer. Based on serum 25(OH)D, vitamin D status was classified as deficiency: <20 ng/ml, insufficiency: 20-29 ng/ml and sufficient: 30–100 ng/ml. [16]

## 3. Statistical analysis

The data was analyzed using Statistical Package of Social science (SPSS) version 11.5 (SPSS Inc., Chicago, USA). We used descriptive statistics to express the baseline variables of the study. Prevalence of vitamin D status across categories of demographic, anthropometric variables and dietary and lifestyle habits were compared using Chi-square test. A p-value of less than 0.05 was considered statistically significant at 95 % confidence interval. Variables found significant in Chi-square test were further subjected to Post-hoc test using Bonferroni corrected p-value. The variables found to be statistically significant were then analyzed by multivariate regression using vitamin D status as the dependent variable.

## 4. Result

### 4.1 Baseline characteristics

Out of 324 participants; 181(55.9%) were female, 102(31.5%) were from Sunsari and 222(68.5%) from Morang. The majority 136(42%) were Brahman and Chhetri. 169(52.2%) were middle-aged and elderly and 96(29.6%) were obese. 164(50.6%) led a sedentary lifestyle. Time of maximum sun exposure for the majority 167(51.5%) was during the late morning. 175(54%) received less than 15 minutes of sun exposure. (Table 1) 305(94.1%) consumed a non-vegetarian diet. 123(38%) consumed mostly chicken. 147(45.4%) consumed meat once a week. Majority 199(61.4%) consumed fish at monthly intervals. 168(51.9%) drank milk less than once a week. 118(36.4%) consumed dairy products 1-2 times/week. (Table 2)

**Table 1:** Baseline characteristics of participants according to vitamin D status.

| Characteristics |  | Total<br>(N=324) |  | Serum vitamin D [25(OH)D] status |  |  |  |  |  | P-value |
| --- | --- | --- | --- | --- | --- | --- | --- | --- | --- | --- |
|  |  |  |  | Deficient<sup>a</sup> |  | Insufficient<sup>b</sup> |  | Sufficient<sup>c</sup> |  |  |
|  |  | N | % | N | % | N | % | N | % |  |
| Age groups | 18-44 years | 155 | 47.8 | 88 | 27.2 | 51 | 15.7 | 16 | 4.9 | 0.1 |
|  | 45-65 years | 169 | 52.2 | 93 | 28.7 | 47 | 14.5 | 29 | 9.0 |  |
| Gender | Male | 143 | 44.1 | 79 | 24.4 | 37 | 11.4 | 27 | 8.3 | 0.04 |
|  | Female | 181 | 55.9 | 102 | 31.5 | 61 | 18.8 | 18 | 5.6 |  |
| Ethnicity | Brahman and Chhetri | 136 | 42.0 | 79 | 24.4 | 37 | 11.4 | 20 | 6.2 | 0.6 |
|  | Newar | 28 | 8.6 | 13 | 4 | 13 | 4 | 2 | 0.6 |  |
|  | Janajati | 130 | 40.1 | 73 | 22.5 | 38 | 11.7 | 19 | 5.9 |  |
|  | Occupational caste (dalit) | 30 | 9.3 | 16 | 4.9 | 10 | 3.1 | 4 | 1.2 |  |
| Region of residence | Sunsari | 102 | 31.5 | 52 | 16.0 | 37 | 11.4 | 13 | 4.0 | 0.2 |
|  | Morang | 222 | 68.5 | 129 | 39.8 | 61 | 18.8 | 32 | 9.9 |  |
| Education | Upto Primary level | 72 | 22.2 | 31 | 9.6 | 24 | 7.4 | 17 | 5.2 | 0.02 |
|  | Intermediate level | 127 | 39.2 | 77 | 23.8 | 32 | 9.9 | 18 | 5.6 |  |
|  | High school | 59 | 18.2 | 33 | 10.2 | 23 | 7.1 | 3 | 0.9 |  |
|  | Above high school | 66 | 20.4 | 40 | 12.3 | 19 | 5.9 | 7 | 2.2 |  |
| Occupation | Professional, SemiProfessional | 54 | 16.7 | 34 | 10.5 | 16 | 4.9 | 4 | 1.2 | 0.4 |
|  | Skilled, Semi-skilled work | 84 | 25.9 | 42 | 13 | 24 | 7.4 | 18 | 5.6 |  |
|  | Arithmetic skill jobs | 42 | 13.0 | 24 | 7.4 | 13 | 4.0 | 5 | 1.5 |  |
|  | Unskilled work | 33 | 10.2 | 22 | 6.8 | 9 | 2.8 | 2 | 0.6 |  |
|  | Unemployed | 111 | 34.3 | 59 | 18.2 | 36 | 11.1 | 16 | 4.9 |  |
| Body Mass Index | Normal | 147 | 45.4 | 80 | 24.7 | 46 | 14.2 | 21 | 6.5 | 0.9 |
|  | Overweight | 81 | 25.0 | 47 | 14.5 | 22 | 6.8 | 12 | 3.7 |  |
|  | Obese | 96 | 29.6 | 54 | 16.7 | 30 | 9.3 | 12 | 3.7 |  |
| Socioeconomic status | Upper middle and above | 83 | 25.6 | 51 | 15.7 | 26 | 8 | 6 | 1.9 | 0.3 |
|  | Lower middle | 112 | 34.6 | 61 | 18.8 | 33 | 10.2 | 18 | 5.6 |  |
|  | Lower class | 129 | 39.8 | 69 | 21.3 | 39 | 12 | 21 | 6.5 |  |
| Smoking Status | Current smoker | 59 | 18.2 | 31 | 9.6 | 17 | 5.2 | 11 | 3.4 | 0.5 |
|  | Former or never smoker | 265 | 81.8 | 150 | 46.3 | 81 | 25 | 34 | 10.5 |  |
| Alcohol intake | Drinks alcoholic beverages | 110 | 34.0 | 64 | 19.8 | 34 | 10.5 | 12 | 3.7 | 0.5 |
|  | Does not drink | 214 | 66.0 | 117 | 36.1 | 64 | 19.8 | 33 | 10.2 |  |
| Physical activities | Moderate | 73 | 22.5 | 40 | 12.3 | 25 | 7.7 | 8 | 2.5 | 0.7 |
|  | Active | 87 | 26.9 | 51 | 15.7 | 25 | 7.7 | 11 | 3.4 |  |
|  | Sedentary | 164 | 50.6 | 90 | 27.8 | 48 | 14.8 | 26 | 8 |  |
| Skin colour | Fair | 92 | 28.4 | 55 | 17.0 | 27 | 8.3 | 10 | 3.1 | 0.001 |
|  | Light brown | 143 | 44.1 | 64 | 19.8 | 57 | 17.6 | 22 | 6.8 |  |
|  | Dark brown | 89 | 27.5 | 62 | 19.1 | 14 | 4.3 | 13 | 4.0 |  |
| Time of maximum sun exposure | Early morning | 108 | 33.3 | 59 | 18.2 | 25 | 7.7 | 24 | 7.4 | 0.02 |
|  | Late morning | 167 | 51.5 | 93 | 28.7 | 58 | 17.9 | 16 | 4.9 |  |
|  | Afternoon | 49 | 15.1 | 29 | 9 | 15 | 4.6 | 5 | 1.5 |  |
| Duration of sun exposure | <15 minutes | 175 | 54.0 | 112 | 34.6 | 43 | 13.3 | 20 | 6.2 | 0.004 |
|  | 15-30 minutes | 86 | 26.5 | 36 | 11.1 | 31 | 9.6 | 19 | 5.9 |  |
|  | >30 minutes | 63 | 19.4 | 33 | 10.2 | 24 | 7.4 | 6 | 1.9 |  |
| Sunscreen | Uses SPF 15 and above | 80 | 24.7 | 41 | 12.7 | 26 | 8.0 | 13 | 4.0 | 0.6 |
|  | Does not use | 244 | 75.3 | 140 | 43.2 | 72 | 22.2 | 32 | 9.9 |  |

Total N% represents the number and percentage of rows.
Deficient<sup>a</sup>: <20 ng/ml, Insufficient<sup>b</sup>: 29 and 20 ng/ml, Sufficient<sup>c</sup>: 30–100 ng/ml 25(OH)D
*P* values <0.05 is significant. *P* value shown in italics was found to be significant on $\chi^2$ test. The *P*-values in bold italics were found to be significant even after post-hoc test using Bonferroni correction.

**Table 2:** Dietary pattern and vitamin D status

| Characteristics |  | Total |  | Serum vitamin D [25(OH)D] status |  |  |  |  |  | *P-value* |
| --- | --- | --- | --- | --- | --- | --- | --- | --- | --- | --- |
|  |  |  |  | Deficient |  | Insufficient |  | Sufficient |  |  |
|  |  | N | % | N | % | N | % | N | % |  |
| Diet | Non-vegetarian | 305 | 94.1 | 171 | 52.8 | 92 | 28.4 | 42 | 13 | 0.8 |
|  | Vegetarian | 19 | 5.9 | 10 | 3.1 | 6 | 1.9 | 3 | 0.9 |  |
| Meat-type | Mostly chicken | 123 | 38.0 | 53 | 16.4 | 44 | 13.6 | 26 | 8 | **0.01** |
|  | Chicken, mutton | 109 | 33.6 | 72 | 22.2 | 26 | 8 | 11 | 3.4 |  |
|  | Chicken, pork | 68 | 21.0 | 43 | 13.3 | 19 | 5.9 | 6 | 1.9 |  |
|  | None | 24 | 7.4 | 13 | 4 | 9 | 2.8 | 2 | 0.6 |  |
| Frequency of meat intake | 2-3 times/month | 50 | 15.4 | 28 | 8.6 | 12 | 3.7 | 10 | 3.1 | 0.6 |
|  | Once a week | 147 | 45.4 | 87 | 26.9 | 42 | 13 | 18 | 5.6 |  |
|  | ≥2/week | 103 | 31.8 | 53 | 16.4 | 35 | 10.8 | 15 | 4.6 |  |
|  | None | 24 | 7.4 | 13 | 4 | 9 | 2.8 | 2 | 0.6 |  |
| Frequency of Fish intake | Monthly | 199 | 61.4 | 109 | 33.6 | 69 | 21.3 | 21 | 6.5 | **0.001** |
|  | weekly | 91 | 28.1 | 45 | 13.9 | 23 | 7.1 | 23 | 7.1 |  |
|  | Rarely/never | 34 | 10.5 | 27 | 8.3 | 6 | 1.9 | 1 | 0.3 |  |
| Frequency of milk intake | Less than once a week | 168 | 51.9 | 101 | 31.2 | 47 | 14.5 | 20 | 6.2 | 0.3 |
|  | 1-2/week | 97 | 29.9 | 52 | 16 | 32 | 9.9 | 13 | 4 |  |
|  | ≥3/week | 59 | 18.2 | 28 | 8.6 | 19 | 5.9 | 12 | 3.7 |  |
| Frequency of dairy products intake | Less than once a week | 115 | 35.5 | 77 | 23.8 | 30 | 9.3 | 8 | 2.5 | **0.02** |
|  | 1-2/week | 118 | 36.4 | 60 | 18.5 | 38 | 11.7 | 20 | 6.2 |  |
|  | ≥3/week | 91 | 28.1 | 44 | 13.6 | 30 | 9.3 | 17 | 5.2 |  |

Total N% represents the number and percentage of rows. *P* value shown in bold italics was significant after post-hoc test using Bonferroni correction.

### 4.2 Serum 25(OH)D status

Overall, 181(55.9%) were vitamin D deficient, 98(30.2%) were insufficient and 45(13.9%) had sufficient levels. (Figure 1) There was significant association between vitamin D status and gender (Chi-sq = 6.2, *p*=0.04), education (Chi-sq = 14.4, *p*=0.02). However, after post-hoc analysis; gender and education did not show significant association with vitamin D. There was significant association between vitamin D status and time of maximum sun exposure (Chi-sq = 11.1, *p*=0.02), duration of sun exposure (Chi-sq = 15.1, *p*=0.004) and skin colour (Chi-sq = 17.9, *p*=0.001). The post-hoc test showed a significantly lower prevalence of vitamin D deficiency among people exposed to sun between 15-30 minutes on an average per day; while getting exposed for > 30 minutes per day had a significantly higher prevalence of deficiency. Similarly, the prevalence of vitamin D sufficiency was significantly higher among people exposed to the early morning sun. Among skin colours, people with dark skin tones had a significantly higher prevalence of vitamin D deficiency than any other skin colour. (Table 1)

**Figure 1:**
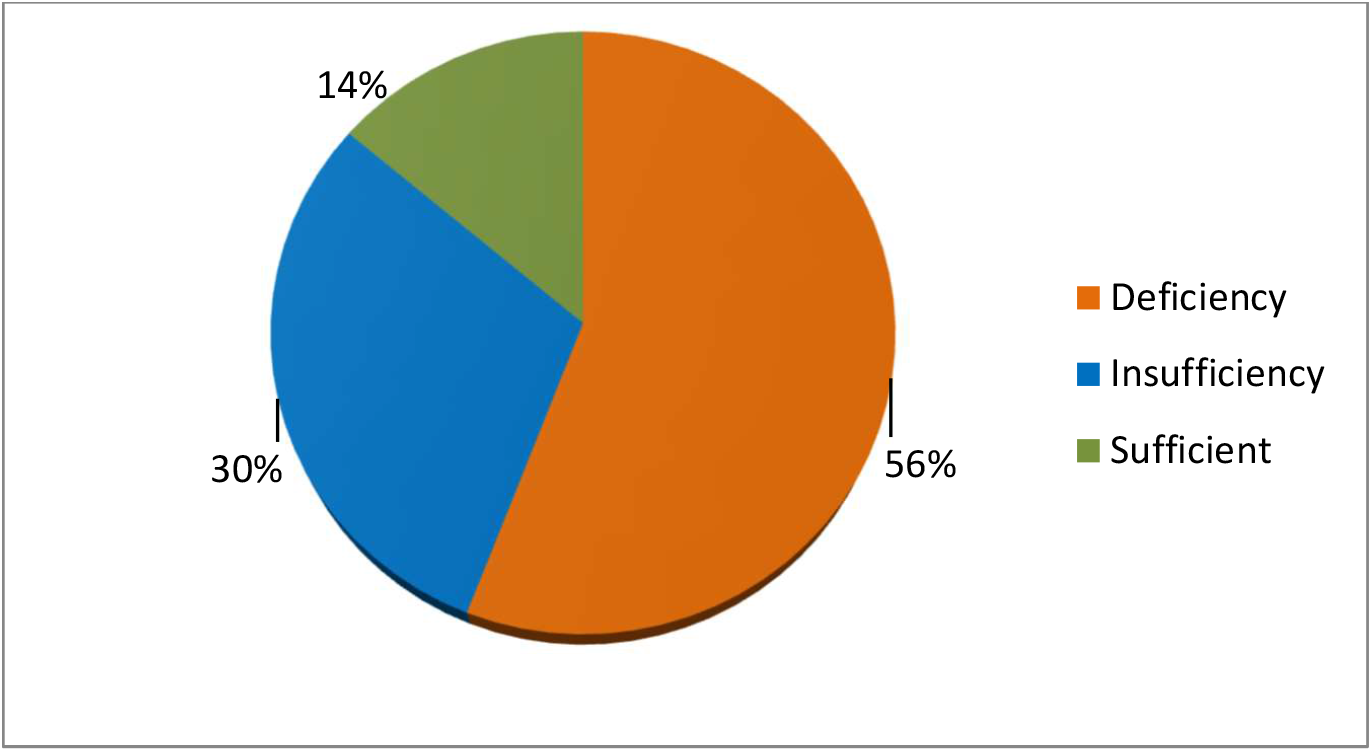
Pie chart showing the distribution of vitamin D status, in Eastern Nepal

Among dietary pattern, we found significant association between vitamin D status and type of meat intake (Fischer’s exact test is 16.4 and p=0.01), frequency of fish intake (Fischer’s exact test is 19.3, p=0.001), frequency of dairy intake (Chi-sq=11.2, *p*=0.02). (Table 2) Post-hoc analysis revealed eating chicken was significantly associated with lower prevalence of vitamin D deficiency; such relation was not established by eating other types of meat. Frequency of fish consumption also showed significant results; people consuming fish weekly had a significantly higher prevalence of vitamin D sufficiency while rarely or never consuming fish had a significantly higher prevalence of deficiency. People consuming dairy products less than once a week had a significantly higher prevalence of vitamin D deficiency.

These characteristics found to be significant on Post-hoc test were further subjected to multivariate logistic regression analysis. The OR (95% CI) for vitamin D deficient and insufficient were compared against the reference vitamin D sufficient. Weekly fish consumption was a significant protective factor with crude OR (95% CI): 0.07(0.009-0.5) *p: 0*.*01* and adjusted OR 0.06(0.008-0.6) *p: 0*.*01*. Consumption of dairy products ≥3/week was also seen as a protective factor against vitamin D deficiency with OR (95% CI): 0.2(0.1-0.6) *p: 0*.*005* and when adjusted with other factors OR was 0.3(0.1-0.8) *p: 0*.*02*. People exposed to the sun for 15-30 minutes per day had lesser odds of vitamin D deficiency than those exposed to lesser duration OR 0.3(0.1-0.7) a *p: 0*.*004*. However, adjusted the odds were not significant. (Table 3)

**Table 3:** Multivariate analysis of vitamin D deficiency and insufficiency.

| Characteristics | Vitamin D status |  |  |  |
| --- | --- | --- | --- | --- |
|  | Vitamin D Deficiency |  | Vitamin D Insufficiency |  |
|  | Crude odds<sup>1</sup><br>(95% CI) | Adjusted odds<sup>2</sup><br>(95% CI) | Crude odds<sup>1</sup><br>(95% CI) | Adjusted odds<sup>2</sup><br>(95% CI) |
| Duration of sun exposure |  |  |  |  |
| >30 minutes | 0.9(0.3-2.6) | 1.2(0.4-3.6) | 1.8(0.6-5.2) | 1.8(0.6-5.6) |
| 15-30 minutes | 0.3(0.1-0.7)<sup>a</sup> | 0.5(0.2-1.1) | 0.7(0.3-1.6) | 1(0.4-2.3) |
| <15 minutes | 1.0 | 1.0 | 1.0 | 1.0 |
| Skin colour |  |  |  |  |
| Fair | 1.1(0.4-2.8) | 0.7(0.3-2.1) | 2.5(0.8-7.1) | 1.6(0.5-4.9) |
| Light brown | 0.6(0.2-1.3) | 0.5(0.2-1.2) | 2.4(0.9-5.9) | 1.8(0.6-4.7) |
| Dark brown | 1.0 | 1.0 | 1.0 | 1.0 |
| Time of max sun exposure |  |  |  |  |
| Early morning | 0.4(0.1-1.2) | 0.5(0.2-1.9) | 0.3(0.1-1.1) | 0.5(0.1-1.9) |
| Late morning | 1.0(0.3-2.9) | 1.3(0.4-4.4) | 1.2(0.3-3.8) | 1.6(0.4-5.4) |
| Afternoon | 1.0 | 1.0 | 1.0 | 1.0 |
| Frequency of dairy intake |  |  |  |  |
| ≥3/week | 0.2(0.1-0.6)<sup>b</sup> | 0.3(0.1-0.8)<sup>c</sup> | 0.4(0.1-1.2) | 0.6(0.1-1.6) |
| 1-2/week | 0.3(0.1-0.7)<sup>d</sup> | 0.33(0.1-0.8)<sup>e</sup> | 0.5(0.1-1.3) | 0.5(0.1-1.4) |
| Less than once a week | 1.0 | 1.0 | 1.0 | 1.0 |
| Frequency of fish intake |  |  |  |  |
| Monthly | 0.1(0.02-1.4) | 0.1(0.01-1.5) | 0.5(0.06-4.8) | 0.8(0.08-7.8) |
| Weekly | 0.07(0.009-0.5)<sup>f</sup> | 0.06(0.008-0.6)<sup>g</sup> | 0.1(0.01-1.4) | 0.2(0.02-2.4) |
| Rarely/never | 1.0 | 1.0 | 1.0 | 1.0 |
| Meat-Type |  |  |  |  |
| Mostly chicken | 0.3(0.06-1.4) | 0.9(0.1-5) | 0.3(0.07-1.8) | 0.6(0.1-3.5) |
| Chicken, mutton | 1(0.2-5) | 3(0.5-18) | 0.5(0.09-2.8) | 1(0.2-6) |
| Chicken, pork | 1.1(0.1-6) | 3.6(0.5-25) | 0.7(0.1-4) | 1.4(0.2-11) |
| None | 1.0 | 1.0 | 1.0 | 1.0 |

Abbreviation: OR Odds ratio, CI: confidence Interval
<sup>1</sup>Univariate logistic regression, <sup>2</sup> Multivariate logistic regression using sufficient vitamin D status as Reference. <sup>a</sup>p-value:0.004 <sup>b</sup>p-value:0.005 <sup>c</sup>p-value:0.02 <sup>d</sup>p-value:0.01 <sup>e</sup>p-value:0.02 <sup>f</sup>p-value:0.01 <sup>g</sup>p-value:0.01

## 5. Discussion

Sunsari and Morang are terai (flat lands) districts of Eastern Nepal with a lower tropical climate. [17] These regions experience abundant sunshine throughout the year. Despite of sunny weather; we found a high prevalence of vitamin D deficiency. We tried to explore different aspects of sun exposure and determine the time of maximum sun exposure and duration of sun exposure needed for optimum vitamin D in this part of the world.

We found people who were exposed to early morning sunlight between 6 am to 8:59 am had significantly greater percentage of sufficient vitamin D levels than those exposed to sun later during the day. This finding contradicts the established notion that sun exposure between 10 am to 3 pm is the best for optimum levels of vitamin D. [5] This may be because most of our participants who were exposed to sun later during the day claimed they used umbrella to shield the hot sun and mostly chose to walk in shady areas while others stated they used transport means to get from one place to another thus decreasing their duration in the sun.

In our study, we determined the optimum duration of sun exposure for adequate vitamin D levels was 15-30 minutes/day. The magnitude of solar radiation reaching the earth’s surface is affected by latitude, season and aerosols. [18] Hence, our finding may be relevant for our latitude, altitude and season of sample collection which were autumn and spring.

We explored if vitamin D status varied between the ethnic groups present in eastern Nepal but found no significant variation. The prevalence of vitamin D deficiency was also comparable amongst the age groups and genders. This may be attributed to the increasing number of women labour force in Nepal venturing outside the confines of the house thus increasing their chances of sun exposure. [19] The gap between the gender further narrows as beauty products as sunscreens are increasingly been marketed to men thus giving both genders equal chances with the sun both in terms of sun exposure and sun barrier.[20]

We found a higher prevalence of vitamin D deficiency among people with dark brown skin. Dark skinned people have larger amounts of melanin pigment which absorbs the ultraviolet rays of the sun and decreases the quantity available for the conversion of 7-dehydrocholesterol to previtamin D3. [8] This suggests that dark skinned people would require greater duration of sun exposure to synthesize sufficient quantity of vitamin D. A study conducted in Northern Europe found insufficient dietary vitamin D and dark skin were the major risk factors for vitamin D insufficiency. [21] Similarly a longitudinal study of parents and children found fairer-skinned children had higher levels of vitamin D. [22]

Dietary sources account for 5 to 10% of the total vitamin D and become crucial where there is inadequate sunlight exposure. [23] Major dietary sources of vitamin D include fish, fish liver oils, beef, pork, chicken, turkey, and eggs. [9] In our study, we analyzed dietary pattern as vegetarian versus non-vegetarian, type of meat consumed, frequency of meat, fish, milk and dairy product consumption. We found participants who consumed fish on weekly basis had significantly higher number of vitamin D sufficient level. Indeed, fish is a natural source of vitamin D proven empirically by a meta-analysis of randomized controlled trial. According to this study consuming ≥2 fish meals/week for at least 4 weeks significantly increased 25(OH)D level. [24] We found people who consumed ≥3times/week dairy products had significantly lower prevalence of vitamin D deficiency. A study conducted to determine the predictors of vitamin D status by Levy MA et al found fortified food, dairy and vitamin D supplement consumption were positive predictors of vitamin D status. [25] Indeed, dairy products as butter and cheese contain up to 10 μg/kg of vitamin D due to its high fat content. [26]

This study has its strengths and limitations. This study helps explore factors determining the vitamin D status in a region of eastern Nepal. It takes into account the various sociocultural factors as ethnic diversity, occupation, dietary habits that are unique to that region of Nepal. However, this paper may also be subject to recall bias as participants answered the questionnaire form based on their recollection. Vitamin D was not measured by HPLC-MS/MS which is considered as reference procedure by International organizations. [27] Hence; our finding may be subject to bias. Furthermore, our study was a cross-sectional one; as a result we cannot establish temporal trajectories.

## 6. Conclusion

The prevalence of vitamin D deficiency was relatively high in eastern Nepal despite adequate sunlight.

## 7. Recommendations

This study highlights the need to create public awareness regarding the importance of vitamin D for good health and the multiple adverse health consequences due to its deficiency. Awareness regarding sun exposure on bare skin for adequate amount of time without using sunscreen should be emphasized. Urbanization and the rush of city life may not spare adequate duration in the sun, in which case nutritional sources and supplements become important. Health organizations and policymakers at regional and national levels need to issue food fortification policies as well as formulate national guidelines for initiating cost-effective regimens for treating vitamin D deficient people.

## Data Availability

All data produced in the present study are available upon reasonable request to the authors

## Author Statement

This study was conducted after receiving ethical approval from Institutional Review Committee, B.P Koirala Institute of Health Sciences (IRC/1220/018).

## Acknowledgement

This research was funded and supported by University Grants Commission, Sanothimi, Bhaktapur, Nepal (UGC Faculty Research Grant for FRG-73/74-HS-04).

